# Factors Influencing the Trajectory of COVID-19 Evolution: A Longitudinal Study of 12 Asian Countries

**DOI:** 10.1101/2023.10.20.23297319

**Authors:** Xiaona He, Hui Liu, Fanyan Zeng, Wei Gao

**Author notes:** Correspondence to: Prof Wei Gao, No. 461, Bayi Ave. Nanchang, Jiangxi China 330006.

## Abstract

**Background:** The effectiveness of different strategies in addressing the COVID-19 pandemic has been assessed, but there is still not enough evidence in Asian countries. This study aims to examine the factors influencing the trajectory of COVID-19 evolution in Asia, to provide insights for optimizing public health policies.

**Methods:** In this longitudinal analysis, we combined COVID-19 cases and vaccination percentages from Our Word in Data with the policy stringency index from the Oxford COVID-19 Government Response Tracker for 12 Asian countries between January 1, 2021, and September 30, 2022. An agglomerative hierarchical cluster analysis (HCA) was conducted to identify countries with similar COVID-19 evolution trajectories. We also investigated the potential impact of seasonal variations on the virus’ trajectory. The relationship between the level of policy response, vaccination coverage, and COVID-19 cases was explored using Generalized Additive Models (GAMs).

**Findings:** There were noticeable differences in the evolution trajectory of COVID-19 among the countries. The 12 Asian countries were grouped into two clusters based on evolutionary similarities. Cluster 1 consisted of West Asian countries (Azerbaijan, Turkey, Bahrain, Israel and Lebanon); while Cluster 2 included Japan, South Korea, Singapore, Malaysia, Thailand, Cambodia and Indonesia. The analysis revealed that the stringency index and vaccination coverage were associated with a statistically significant impact (both *P* values < 0·0001) on the evolution trajectory of COVID-19 (_adj_R^2^=0·54). The dose-response relationships demonstrated that the continuous high levels of stringency index (≥87·6) or vaccination coverage (≥ 42·0%) have led to a decrease in COVID-19 infection rates. In early 2021, the _adj_R^2^ increased to 0·93 for all countries. Furthermore, the _adj_R^2^ for Cluster 1 and Cluster 2 were 0·86 and 0·90 respectively. All GAMs models have significantly improved compared to null model (*P* values <0·0001).

**Interpretation:** By strengthening vaccination ahead of susceptible seasons and enhancing personal self-protection measures, the transmission of COVID-19 among the population can be reduced even during the highly infectious Omicron era.

**Funding:** Senior Talent Startup Fund of Nanchang University

## Introduction

Asia is the most populous continent globally, housing over 4·7 billion people, which accounts for 59·3% of the global population. ^1^ Due to its large population size and high population density, the COVID-19 pandemic has posed significant challenges for countries in Asia. Consequently, these countries have implemented various strategies to combat the pandemic.

Interestingly, there are notable differences in the progression of COVID-19 among Asian countries. The emergence and spread of variants of concern (VOCs), such as Alpha, Beta, Gamma, and Delta, ^2^ have led to a global upsurge of infections in 2021. Some Asian countries, such as Singapore, South Korea, Japan, ^3^ and Cambodia, ^4^ have consistently maintained a relatively low and stable level of infections. In contrast, countries like Israel, Bahrain and Malaysia ^5^ have experienced fluctuations, with significant increases in the number of cases. These variations may be attributed to differences in countries’ responses to COVID-19, ^6–8^ including government policy measures ^9^ and the implementation of vaccination programs. ^10^ However, with the emergence of the highly transmissible Omicron variant and its global dominance starting in January 2022, ^11–13^ the situation has significantly shifted. The destructive nature of the Omicron variant, coupled with public fatigue towards long-term containment measures, has led many Asian countries to relax non-pharmaceutical interventions (NPIs). As a result, certain countries had witnessed unprecedented increases in infection cases. For instance, South Korea,^14^ which maintained relatively lower infection levels in 2021, experienced a rapid surge in daily COVID-19 cases in early 2022.

Understanding the epidemiological characteristics of infectious diseases is crucial for controlling their transmission. Gaining insights into the evolving dynamics of COVID-19 and its influencing factors within Asian countries can provide valuable information for future prevention and control strategies. And yet, there is still a lack of enough research on this subject in various Asian countries over an extended period. Hence, our study sought to fill this knowledge gap by examining and contrasting the COVID-19 progressions in different Asian nations. Additionally, we aimed to investigate the influence of seasonal fluctuations, government policy response, and vaccination coverage on the development of COVID-19 trajectories.

## Methods

### Data sources and variable definitions

We extracted the seven-day moving average of daily cases per million and the percentage of people fully vaccinated from Our Word in Data. ^15^ Additionally, we obtained the government response stringency index, which is a comprehensive indicator of government response, from the Oxford COVID-19 Government Response Tracker (OxCGRT).

1. 7-day moving average of daily cases per million: this measure is commonly used by policymakers to assess the spread of COVID-19. It helps to smooth out fluctuations in daily cases that may arise from reporting or counting errors, rather than actual daily cases. We used the number of the 7-day moving average of daily cases per million to represent the infection rate;
2. Percentage of people fully vaccinated: this refers to the total number of people who have received all the doses prescribed by the initial vaccination protocol. We used this variable to represent vaccination coverage. Considering the sufficient antibodies in the body after taking the second vaccine dose, ^16^ we used the value of this indicator with a seven-day lag time to analyze its impact on COVID-19 evolution.
3. Stringency index : this index is used to reflect the level of policy response. ^17^ It is a composite measure based on nine response indicators, including school closures, workplace closures, cancellation of public events, restrictions on gathering size, closure of public transport, stay-at-home requirements, restrictions on internal movement, restrictions on international travel and public information campaign. The index is scored from 0 to 100, with a higher score indicating a higher level of policy response. We used a seven-day lag time of the stringency index to account for the delay effect for response measures in controlling the spread of COVID-19. It is important to note that the stringency index does not measure the effectiveness of government policies in response to COVID-19, but rather the extent and comprehensiveness of governmental response.
4. Country specific variables: we also considered some variables that may directly influence COVID-19 infection rates, including population density, urban population (% of total population) and particulate matter 2.5 (PM 2.5) air pollution mean annual exposure. These variables represent national characteristics before the emergence of COVID-19 and were obtained from the World Bank in 2019.

### Study period

Taking into consideration the background of vaccination and the start of vaccination efforts in most Asian countries in 2021, along with the relaxation of policies and the subsequent discontinuation of strict case reporting in late 2022, we selected the research period from January 1, 2021, to September 30, 2022 to ensure reliability of the data.

To analyze the unique spread pattern of the Omicron-dominant variant, we divided the data into two separate periods. Period 1 represented the time before Omicron became predominant (January 1, 2021 - December 31, 2021), while Period 2 represented the period when Omicron was the dominant variant (January 1, 2022 - September 30, 2022).

### Data preparation

To ensure the reliability of the data, we checked for missing and incorrect data. Since different countries started their vaccination programs at different times and some did not publish daily coverage data, we excluded countries with a substantial amount (≥50%) of missing data. The final sample consisted of 12 countries: Azerbaijan, Turkey, Bahrain, Israel, Lebanon, Japan, South Korea, Singapore, Malaysia, Thailand, Cambodia, and Indonesia.

For the few missing values within these 12 countries, our approach was as follows: (1) when there were missing values corresponding to the time before the country started its second dose vaccination program, those missing values were defined as 0; (2) for the small proportion of missing values that occurred during the country’s vaccination program, we used linear interpolation to impute the missing values.

### Statistical analysis

We described the evolution trajectory of COVID-19 from January 1, 2021 to September 30, 2022. To identify countries with similar trends, we performed an agglomerative hierarchical cluster analysis (HCA).^20^ We used Pearson correlation-based distance as a measure of dissimilarity. This allowed us to identify clusters of observations with similar overall trends. We then applied Ward’s method to measure the distance between clusters. Before conducting HCA, we normalize the original data using z-score transformation, resulting in a mean of 0 and a variance of 1.

We also considered the potential influence of the seasonal cycle on the evolution trajectory of COVID-19. To analyze the seasonal variations in the normalize number of daily confirmed cases, we employed the Complete Ensemble Empirical Mode Decomposition with Adaptive Noise (CEEMDAN) method.^18^ This method is used to decompose and analyze time series data that may be nonlinear and/or non-stationary.

To investigate the impact of the level of policy stringency index and vaccination coverage on the evolution trajectory of COVID-19, we used generalized additive models (GAMs). These models were employed to examine the smooth functional relationships between the average weekly level of policy response, vaccination rate, and the cumulative number of weekly COVID-19 cases. The GAMs were primarily conducted during Period 1, which was the main period for government policy response and vaccination. GAMs are a combination of the generalized linear model (GLM) and the additive model (AM). ^19^ These data-driven non-parametric regression models offer the flexibility to capture complex relationships by considering non-linear effects. The formula of the model was defined as follows:

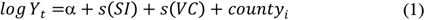

In the formula, log (*Y*_*t*_) represents the logarithm of the cumulative number of weekly COVID-19 cases; *α* is the intercept; *SI* represents the weekly average level of policy response (stringency index) to COVID-19; *VC* represents the weekly average vaccination rate foe COVID-19; *s ()* refers to a thin plate spline function based on the penalized smoothing spline. Each additive term in the GAMs is estimated using a single smooth function, which explains how the dependent variable changes with the corresponding independent variable. *Country*_*i*_ is a dummy variable that represents each country and controls for country-level characteristics. The GAMs use a Gaussian distribution family and are fitted using restricted maximum likelihood (REML) ^20^ to estimate model coefficients and smoothing parameters.

All analyses were performed using R software (version 4.3.1). The pheatmap ^21^ package was used to implement HCA, the Rlibeemd package ^22^ was employed to extract seasonal signals and the mgcv package^23^ was used to fit the GAMs, and the A significance level of *P* < 0·05 was considered indicative of statistically significant.

### Role of the funding source

The funder of the study had no role in study design, data collection, data analysis, data interpretation, or writing of the report.

## Results

### The evolution trajectory of COVID-19 and country profiles in 12 Asia countries

Figure 1 shows the evolution trajectories of COVID-19 among 12 Asia countries. During Period 1, there were significant differences in the evolution trajectories of COVID-19 across these countries. In the first half of 2021, several countries experienced a surge in confirmed COVID-19 cases. Israel, Lebanon and Bahrain witnessed the onset of their first wave in early 2021, with the highest daily confirmed cases of COVID-19 per 100,000 being 912·7, 874·5, 2021·0 respectively. Turkey and Azerbaijan began to observe an increase in cases starting in April, peaking at 706·2 and 216·9, respectively. In contrast, countries like Singapore, South Korea and Japan maintained a steady low confirmed COVID-19 cases, with the highest at 7·0, 18·4 and 52·4, respectively. By the second half of 2021, a rise in confirmed COVID-19 cases was observe in most countries. The timing and intensity of these peaks varied among nations. Malaysia reached its peak in August, while Israel in early September and Singapore at the end of October.

**Figure 1.**
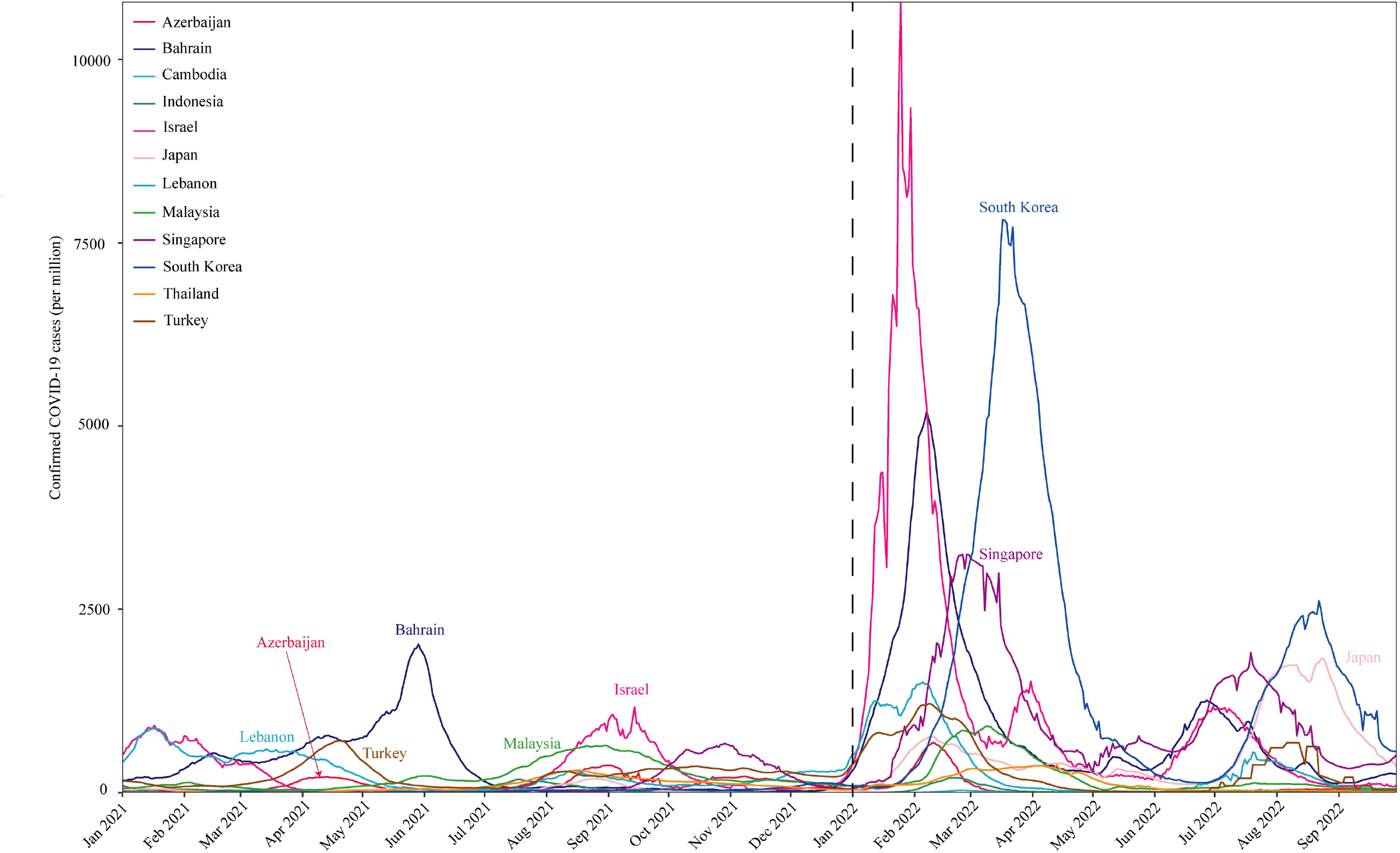
The trajectories of daily confirmed COVID-19 cases per million among the 12 countries, Jan, 2021-Sep, 2022. The vertical dotted line is the dividing line between the two phases (on the left is period 1, on the right is period 2).

In Period 2, starting from January 2022, the majority of countries experienced a sharp increase in confirmed COVID-19 cases. Furthermore, most countries faced two waves of pandemic between January and September 2022. The first wave occurred primarily from January to March, while the second wave predominantly took place from July to September.

Table 1 provides the characteristics for each country. Prior to Period 2, Bahrain had the highest number of cumulative confirmed cases per 100,000 (12943), followed by Israel (10272) and Lebanon (9770). On the other hand, Cambodia had the lowest number of cases (724), followed by South Korea (1108) and Japan (1192). During Period 2, South Korea had the highest number of cumulative confirmed cases per 100,000 (46692), followed by Israel (35013), Singapore (29861), Bahrain (27233) and Japan (27233). Cambodia (105) and Indonesia (792) had the lowest number of cumulative confirmed cases per 100,000. The top six countries with the highest population density were Singapore, Bahrain, Lebanon, South Korea, Israel and Japan. Additionally, these countries also had the largest urban population as a percentage of the total population. On the other hand, Cambodia, Thailand, Indonesia and Azerbaijan had low population density and urban population (% of total population). The country with the lowest PM2·5 was Japan (13), followed by Malaysia (17), Singapore (19) and Indonesia (19). In contrast, Bahrain recorded the highest PM2·5(59).

**Table 1.** The characteristics for the 12 Asian countries.

| Countries | Cumulative number of cases per 100,000 (Period 1) | Urban population (% of total population) | Population density | PM 2.5 | Cumulative number of cases per 100,000 (Period 2) |
| --- | --- | --- | --- | --- | --- |
| Cambodia | 724 | 24 | 92 | 22 | 105 |
| South Korea | 1108 | 81 | 530 | 27 | 46692 |
| Japan | 1192 | 92 | 347 | 13 | 15564 |
| Indonesia | 1286 | 56 | 144 | 19 | 792 |
| Thailand | 3096 | 51 | 140 | 27 | 3447 |
| Azerbaijan | 3928 | 56 | 121 | 25 | 2017 |
| Singapore | 4049 | 100 | 7966 | 19 | 29861 |
| Malaysia | 7878 | 77 | 100 | 17 | 6204 |
| Turkey | 8580 | 76 | 108 | 26 | 8719 |
| Lebanon | 9770 | 89 | 565 | 29 | 8724 |
| Israel | 10272 | 93 | 418 | 20 | 35013 |
| Bahrain | 12943 | 89 | 1908 | 59 | 27233 |

Note: The cumulative numbers of infections in Period 1 and Period 2 aren't directly compared, as Period 1 contains twelve months of data, while Period 2 only contains nine months.

### Similar trends in the evolution trajectory of COVID-19

Figure 2 shows that the 12 Asian countries can be divided into 2 main clusters based on their similarities in evolution. Cluster 1 consists of five West Asian countries (Azerbaijan, Turkey, Bahrain, Israel and Lebanon). Cluster 2 includes two developed countries (Japan and South Korea) in East Asia, one developed country (Singapore) in Southeast Asia, and four Southeast Asian countries (Malaysia, Thailand, Cambodia and Indonesia). The dendrogram in Figure shows that Azerbaijan and Turkey cluster together, indicating a high degree of similarity between them. Similarly, Bahrain, Israel and Lebanon cluster together due to their comparable evolution trajectories, with Israel and Lebanon exhibiting a closer evolution trajectory compared to Bahrain. Japan, Singapore and South Korea share the same grouping, while Malaysia and Thailand form another cluster. Cambodia and Indonesia also share a common grouping. It is worth noting that, due to the predominance of the Omicron variant, countries in Cluster 2 experienced their first wave of peaks between February and March. Conversely, for countries in Cluster 1, the peak of the initial wave of the epidemic in 2022 came earlier, from January to February.

**Figure 2.**
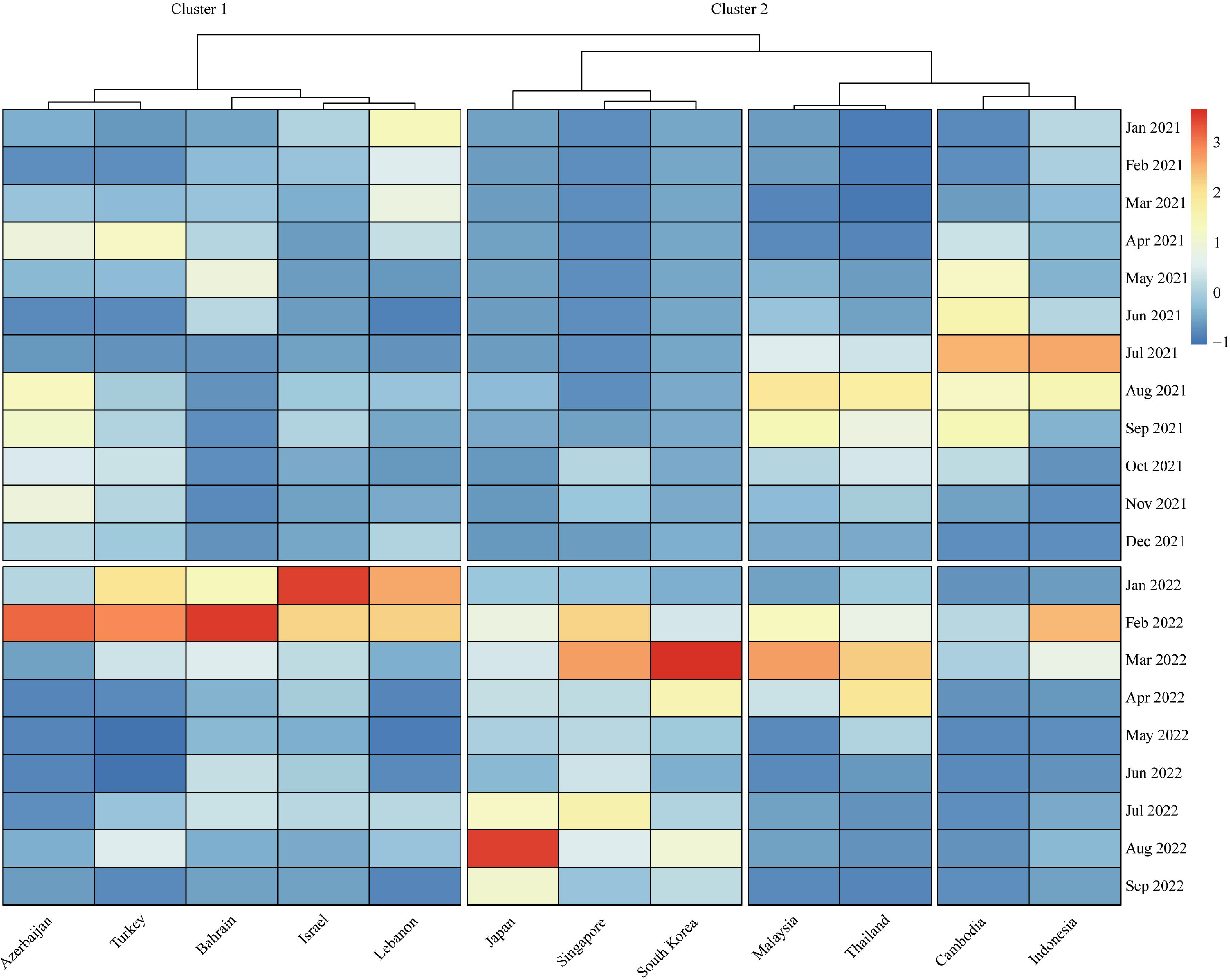
Heat-map representation of confirmed COVID-19 cases in the 12 countries, Jan, 2021-Sep, 2022. The height of nodes connecting two samples on the dendrogram represents the degree of dissimilarity. When the connection node is lower, it indicates a higher the degree of similarity between samples. For example, Singapore is more similar to South Korea than it is to Japan. The color represents the Z-score transformation value of the original data of the country, the larger the value, the larger the number of cases relative to other months in the country.

Moreover, countries with similar evolution trajectories of COVID-19 also displayed comparable patterns in terms of the stringency index. Figure 3 illustrates the countries with similar evolution trajectories and their corresponding stringency index. These countries differ in the strictness of responses towards the COVID-19 pandemic. As of March 31, 2021, Israel (50·9%) had the highest vaccination coverage, followed by Bahrain (17·4), Turkey (8·0%), and Singapore (7·2%). The remaining countries had relatively low vaccination coverage (<2·0%). Supplementary File Figure S1 provides information on the vaccination coverage trends for each country.

**Figure 3.**
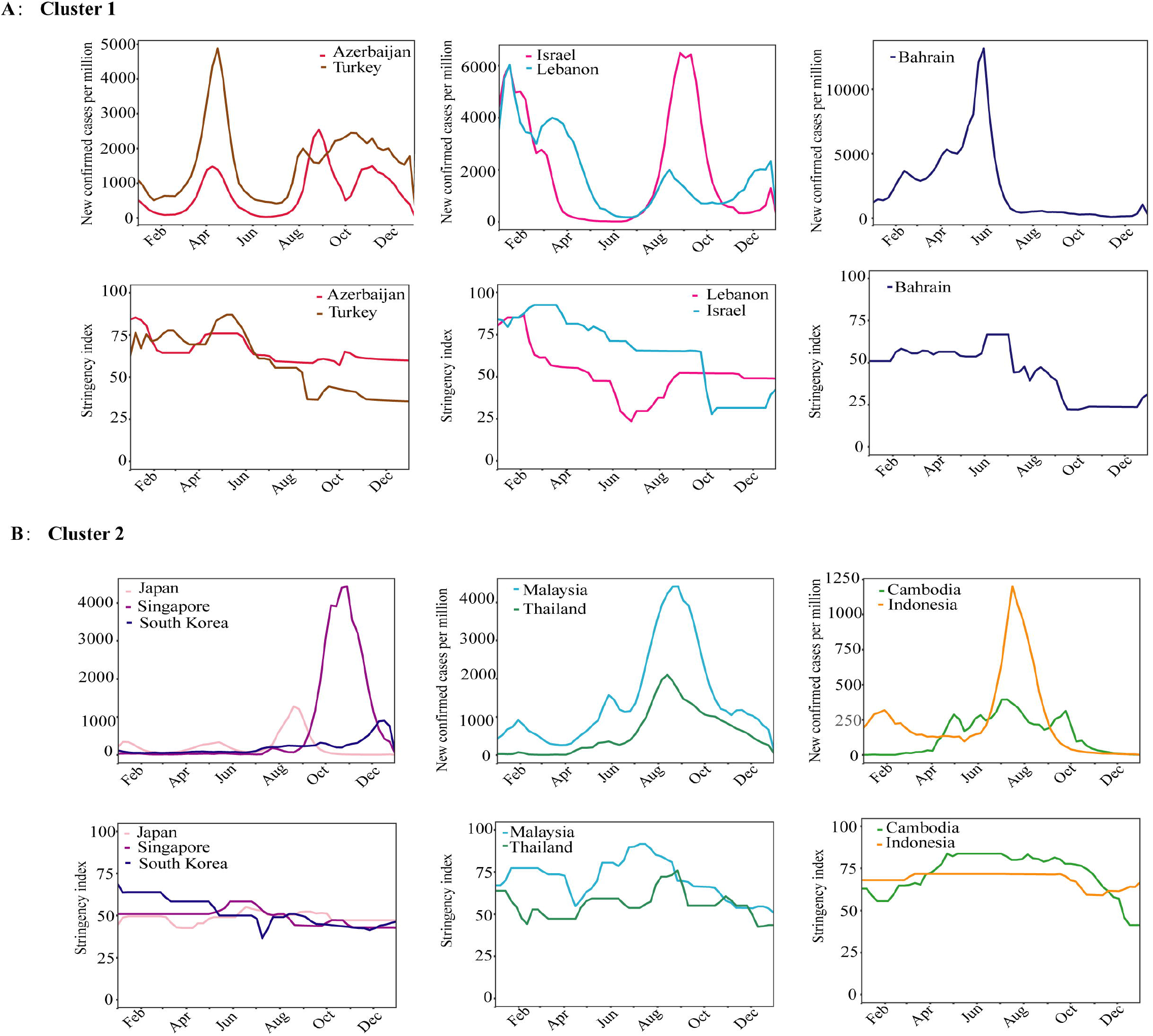
Countries with similar evolution trajectories and their stringency indices from January 2021 to December 2021. A) Cluster 1; B) Cluster 2.

**Figure 4.**
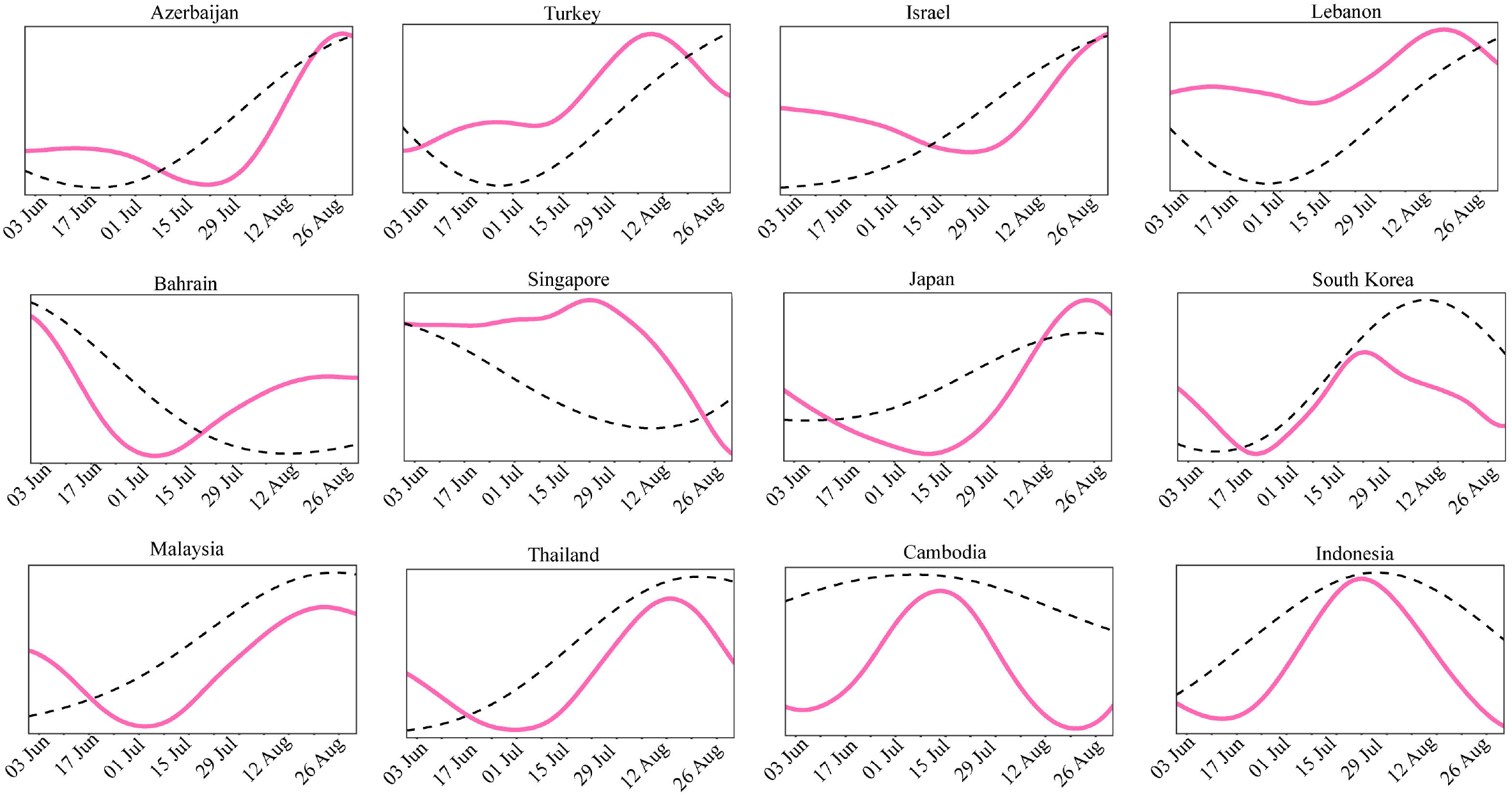
The seasonal components extracted from daily COVID-19 confirmed cases with CEEMDAN for 12 Asia countries in June to August 2021. The black dashed line represent the overall trend. The pink solid line represent the seasonal component. A) Seasonal components of Cluster1 including 4 West Asian countries; B) Seasonal components of Cluster2 including Japan, Singapore, South Korea and 4 other Southeast Asian countries.

### Seasonal variations in the evolution trajectory of COVID-19

We observed similar seasonal fluctuations among these 12 countries from January 2021 to September 2022, especially during the summer months, with July showing the most distinct pattern. According to Figure 3, the seasonal composition for the 12 countries during the summer of 2021 indicated that, except for Azerbaijan, which experienced a rise in cases starting from August 3rd, all other countries showed an upward trend in confirmed cases in July. Furthermore, Figure S2-S3 depicted the seasonal variations for the 12 countries from January 2021 to September 2022.

### The relationship between the level of stringency index, vaccination coverage, and the evolution trajectory of COVID-19

Equation (1) was used to investigate the impact of the level of stringency index and vaccination coverage on the evolution trajectory of COVID-19. We choose the model based on the principle of minimum Akaike information criterion (AIC), and the specific values can be found in Supplementary Table S1. The results indicated that both the level of stringency index and vaccination coverage had a statistically significant impact (both *P* values <0·0001) on the evolution trajectory of COVID-19, with an adjusted R^2^ of 0·54. The adjusted R^2^ reflects the degree of fit of the model. Compared to null model(_adj_R^2^=0·37), the full model has significant improvements (*P* value <0·0001).

The GAM dose-response curve (Figure 5A) suggested that increasing the level of stringency index could help control the spread of infection as the number of infections increase. A noticeable reduction in the number of infections was observed higher than strict level of 87·6. Additionally, a vaccine coverage rate of higher than 42·0% contributed to reducing the number of infections (Figure 5A).

**Figure 5.**
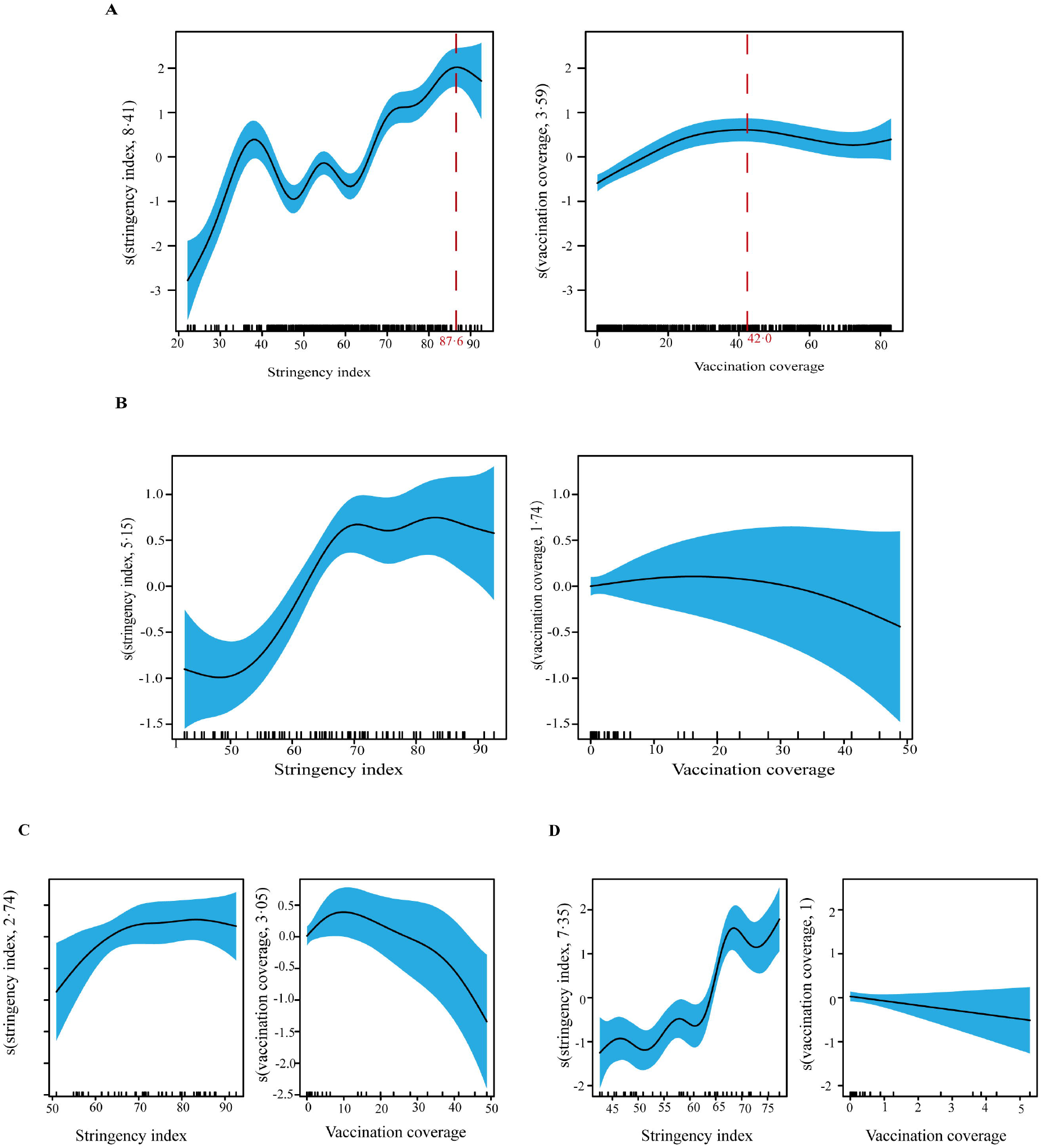
Exposure–response curves on the effects of the stringency index and vaccination coverage in COVID-19 cases. The x-axis represents independent variable (stringency index (on the left panel) and vaccination coverage (right panel)). The y-axis shows the contribution of the smoother to the fitted values. The black line is the expected value of the effect, shaded blue is 95% confidence bands for smooths. A) The 12 countries from January 2021 to December 2021; B) The 12 countries from January 2021 to March 2021; C) Cluster1 including 5 West Asian countries(Azerbaijan, Turkey, Bahrain, Israel and Lebanon) from January 2021 to March 2021; D) Cluster2 including Japan, Singapore, South Korea and 4 other Southeast Asian countries(Malaysia, Thailand, Cambodia and Indonesia) from January 2021 to March 2021. Need to pay attention to the difference in vertical coordinates.

In addition, when urban population (% of total population) and PM2·5 were included as covariates in the GAM model, it was found that both variables were positively correlated with the infection level (*β*_1_=0·04, *β*_2_=0·15 respectively, and *P* value <0·0001). However, they did not have a significant impact on the model, as the adjusted R^2^ remained at 0·54.

We conducted a segmental time analysis for the period from January to March 2021 to evaluate the potential influence of seasonal fluctuations. It was observed that the sustained high levels of the stringency index and vaccination resulted in a reduction in COVID-19 infections (Figure 5B). The adjusted R^2^ of the model increased to 0·93, indicating a strong correlation. Compared to null model(_adj_R^2^=0·91), the full model has significant improvements (*P* value <0·0001).

Furthermore, separate GAMs were constructed for the two categories of countries. It was found that the policy stringency index varied between the countries in Cluster 1 and Cluster 2 during this period. Despite this variation, both clusters demonstrated a significant impact in lowering infection levels through rapid vaccine administration, as illustrated in Figure 5C-D. The adjusted R^2^ for Cluster 1 and Cluster 2 were 0·86 and 0·90, respectively. The corresponding null models _adj_R^2^ were 0·75 and 0·83, respectively, and both full models had significantly improved(*P*<0·0001). Furthermore, it is worth noting that there was a linear trend in the vaccination coverage for Cluster 2, with a linear regression coefficient of *β* = -0·12(*P*=0·09).

### Sensitivity Analysis

We conducted cluster analysis using the daily and weekly cumulative number of confirmed cases, which yielded consistent classification results. We also adjusted the calculation of the distance matrix by applying the Euclidean distance. However, the classification results remained unchanged. The results of this analysis are presented in Figure S4-S6.

In the fitted GAM model, we performed model checking, and the results suggested that our model had a good fitting effect. The results were presented in Figure S7. In addition, the collinearity of the fitted GAM model was tested using the ‘concurvity’ function from the ‘mgcv’ package in R, based on the variance inflation factor (VIF). For linear calibration, the VIF values of the explanatory variables were found to be less than 5.

We also examined the stringency index and vaccination coverage without considering lag effects and with a 14-day lag in the GAM model. Interestingly, we arrived at similar conclusions in both scenarios shown in Figure S8 and Figure S9. Furthermore, we examined the impact of government policy response and vaccination during Period 2 which was the period dominated by Omicron variant. The results of the analysis supported our key research findings. This analysis is shown in the Figure S10-S11 of the Supplementary file.

## Discussion

This study presents the first comprehensive analysis of the factors influencing the trajectory of COVID-19 evolution in Asia. Our study reveals the significant role played by the level of policy response and vaccination coverage in shaping the evolution trajectory of COVID-19.

We observed a decline in the epidemic curve in countries that consistently implemented a high level of stringency index and rapidly expanded vaccination campaigns during the initial wave of the COVID-19 pandemic in early 2021. For instance, Israel, which maintained an average stringency index of 85·5 and rapidly rolled out mass vaccination, witnessed a significant reduction in new infection cases in January 2021. This observation was supported by previous findings. ^24^ Similarly, Lebanon followed a similar epidemic trajectory to Israel, gradually reducing infection cases by relying on sustained strict interventions in the absence of rapid vaccine distribution. In contrast, Bahrain, despite experiencing a continuous increase in new infection cases, implemented a lower level of stringency index (average SI=52·3) during the same period. Previous research has also identified the relaxation of outdoor activity restrictions in Bahrain as a contributing factor to the outbreak and increased number of COVID-19 cases. ^25^

Countries like Japan, Singapore, and South Korea, which consistently maintained a moderate level of stringency index, also exhibited low infection rates. With the exception of Singapore, which rapidly expanded vaccination in February 2021, Japan and South Korea experienced a delayed implementation of the second dose of vaccines. We found that the sustained moderate level of stringency index in these countries contributed to maintaining low infection rates. Previous findings suggest that the effectiveness of social distancing measures in these countries along with high compliance among residents played a significant role ^26^.

Seasonality is another important factor that affects the evolution trajectory of COVID-19. We are particularly interested in the summer season, as these 12 countries have observed a consistent surge in cases during this time. Despite the Summer Olympics taking place in Tokyo, Japan on July 23, 2021, a study showed that the increase in COVID-19 cases during that period had little to do with the event. ^27^ Apart from seasonal fluctuations, other studies have indicated that the spread of Delta variants has contributed to the impact of the summer surge in 2021. ^28,29^ Nonetheless, during the dominant period of Omicron variants, in addition to the initial peak from January to March 2022, a second peak has also emerged in the summer. A study analyzing wastewater in Bahrain revealed that during the summer of 2022, the positivity rate of SARS-CoV-2 virus in the wastewater was notably high, ranking second only to the rate recorded in January. ^30^ Therefore, it is important to pay attention to the fluctuations of COVID-19 during the summer.

Our research has some limitations. Firstly, we did not take into account the proportion of different variants in each country. Instead, we defined two periods based on the unique transmission patterns of the Omicron variants and conducted grouped analyses considering countries with similar geographical locations and the evolution trajectory of COVID-19. This partially compensates for this limitation. Secondly, due to reporting deficiencies, the number of COVID-19 infections might be underestimated, which may require a more conservative interpretation of the results. Third, we analyzed at the country level, so changes at smaller unit levels, such as provincial, community levels, may have been overlooked. Additionally, the evidence presented in this article cannot prove a causal relationship between these factors and COVID-19 outcomes.

This analysis provides a comprehensive assessment of the factors influencing the trajectory of COVID-19 evolution in Asia. The findings obtained from this study could offer insights for optimizing public health policies.

## Data Availability

All data produced are available online at Our World in Data and the Oxford COVID-19 Government Response Tracker

https://ourworldindata.org/coronavirus

https://www.bsg.ox.ac.uk/research/covid-19-government-response-tracker

## Contributors

WG conceived the research idea, designed and supervised the study. XH performed data collection, management and analysis. HL and FZ contributed to the collection and curation of data. XH drafted the manuscript. WG directed the study, and critically revised the manuscript. All authors had full access to all the data in the study and verified the data. WG had final responsibility for the decision to submit for publication.

## Declaration of interests

We declare no competing interests.

## Data sharing

All data are publicly available and listed in this Article.

## Acknowledgments

We thank Our World in Data and the Oxford COVID-19 Government Response Tracker project for providing free and accessible data, as well as to the volunteer coders involved in both projects.

## Notes

### Competing Interest Statement

The authors have declared no competing interest.

### Funding Statement

This study was funded by Senior Talent Startup Fund of Nanchang University

